# Toward noninvasive assessment of stroke risk in pediatric cerebrovascular disease

**DOI:** 10.1101/2021.12.17.21267944

**Authors:** John D. Horn, Michael J. Johnson, Zbigniew Starosolski, Avner Meoded, Dianna M. Milewicz, Ananth Annapragada, Shaolie S. Hossain

## Abstract

**Background and Purpose:** Moyamoya disease (MMD) is a progressive steno-occlusive cerebrovascular disease leading to recurrent stroke. There is a lack of reliable biomarkers to identify unilateral stroke MMD patients who are likely to progress to bilateral disease and experience subsequent contralateral stroke(s). We hypothesized that local hemodynamics are predictive of future stroke and set out to noninvasively assess this stroke risk in pediatric MMD patients.

**Methods:** MR and X-ray angiography imaging were utilized to reconstruct patient-specific models of the circle of Willis of 6 pediatric MMD patients who had previous strokes, along with a control subject. Blood flow simulations were performed by using a Navier-Stokes solver within an isogeometric analysis framework. Vascular regions with a wall shear rate (WSR) above the coagulation limit (> 5000 s^−1^) were identified to have a higher probability of thrombus formation, potentially leading to ischemic stroke(s). Two metrics, namely, “critical WSR coverage” and “WSR score”, were derived to assess contralateral stroke risk and compared with clinical follow-up data.

**Results:** In two patients that suffered a contralateral stroke within two months of the primary stroke, critical WSR coverages exceeding 50% of vessel surface and WSR scores greater than 6x the control were present in multiple contralateral vessels. These metrics were not as conclusive in two additional patients with 3-to-5-year gaps between primary and contralateral strokes. However, a longitudinal study of one of these two cases, where a subsequent timepoint was analyzed, accurately predicted disease stabilization on the primary stroke side and an elevated contralateral stroke risk, thus indicating that post-stroke follow-up at regular intervals might be warranted for secondary stroke prevention.

**Conclusions:** WSR-based metrics could be predictive of future stroke risk after an initial stroke in MMD patients. In addition, more accurate predictions may be possible by performing patient-specific hemodynamic analysis at multiple timepoints during patient follow-up to monitor changes in the WSR-based metrics.

## 1. Introduction

Moyamoya disease (MMD) is a progressive cerebrovascular disorder characterized by stenotic or occlusive lesions in the terminal internal carotid artery (ICA) and the proximal middle or anterior cerebral arteries (MCA, ACA), leading to recurrent transient ischemic attacks or stroke. A substantial number of patients presenting with unilateral lesions go on to form bilateral disease, and it is known to occur more commonly in pediatric patients^1^. Neurosurgical intervention on the symptomatic arterial lesion, followed by close observation for bilateral involvement is usually recommended^2^, as unilateral stroke is sometimes followed by contralateral stroke(s), even after receiving surgical treatment^3^. However, there is no reliable way to identify at-risk patients and intervene before the subsequent stroke occurs. Progression of vasculopathy associated with MMD is highly variable from patient to patient, and patients with similar angiographic appearance can have varying degree of stroke risk^4^. As a result, several studies that have relied on angiographic appearance to predict progression to bilateral disease, produced conflicting conclusions^5^.

While pathophysiology of MMD remain unclear, hemodynamic stress is known to play an important role in stroke^4, 6^. A vascular wall shear rate (WSR) above the coagulation limit (> 5000 s^−1^) signifies an increased probability of thrombus formation^7^, leading to stroke^8^. In a preliminary study using an *Acta2*^*-/-*^ mouse model that develops many of the features of MMD^9, 10^ we demonstrated that occlusion in one of the major arteries in the circle of Willis (CoW) increased hemodynamic burden in vessels contralateral to the occlusion, thus potentially raising contralateral stroke risk if critical WSR limits are exceeded^3^. If a similar behavior is realized in the MMD patients, it could have a profound implication for patient care. We therefore hypothesized that local WSR could be predictive of future stroke and that image-based patient-specific hemodynamic analysis, in conjunction with clinical observations, could be used to noninvasively assess this stroke risk in pediatric MMD patients.

In a recent study, Lee et al. hypothesized that endothelial shear stress could act as a trigger for alteration of vascular remodeling and correlated contralateral progression of unilateral MMD to spatial variability of endothelial shear stress around contralateral terminal ICA^11^. This wall shear stress was indirectly estimated by using signal intensity gradient (SIG) in time-of-flight (TOF) sequences from brain MRI, but validation with computational fluid dynamics (CFD) was not performed. In the present work, we conduct physiologically realistic CFD analysis of the full CoW and directly quantify local WSR distribution for a pilot cohort (n = 6) of pediatric MMD patients. The objective is to identify susceptible vascular regions with WSR critically above 5000 s^−1^ that could evolve into severe stenosis or complete occlusion due to thrombus formation, leading to ischemic strokes^8^. The in-silico findings are compared with patients’ outcome data to investigate whether such subject-specific analysis of local WSR utilizing patient’s post-stroke MR TOF and X-ray angiography (XA) images is predictive of future stroke. The findings of this study could inform follow-up strategies for unilateral patients at risk for contralateral stroke and potentially guide timing of preventive surgical interventions.

## 2. Methods

### 2.1 Vascular model creation from imaging data

The generation of patient-specific CoW models from MR TOF images and their adjustment with XA data are outlined below.

#### 2.1.1 MR TOF segmentation

Patient MR TOF images (**Supplementary Figure S1A**) are segmented in 3D Slicer using a combination of intensity thresholding and manually painting vessels in each slice. The final segmented label map is exported as a triangulated 3D surface mesh and is considered the initial model (**Supplementary Figure S1B**).

#### 2.1.2 Diameter adjustment using XA images

3D MR TOF images can have poor signal where blood flow is slow or complex, causing vessel caliber to be inaccurately represented^12^. Inadequate image resolution can cause areas of MMD-related severe narrowing to be poorly resolved, resulting in inaccurate evaluation of local hemodynamics^3^. To improve accuracy of vessel geometry (e.g., vessel diameter), the MR-derived CoW model (initial model) is adjusted based on 2D imaging data obtained through XA (**Supplementary Figure S1C**), which is considered the gold standard for evaluating vessel narrowing.

Details of this method can be found elsewhere (BIORXIV/2021/472309). Briefly, the initial model is first aligned to the XA imaging view to enable comparison of the initial CoW model vessel geometry with that seen on 2D XA images. For each XA image, an imaging axis is defined in a virtual space by using XA imaging metadata, including *distance source to patient, distance source to detector, positioner primary angle*, and *positioner secondary angle*. Centerlines are extracted from the initial model and projected along the imaging axis onto the virtual detector plane. Using custom algorithms implemented in MATLAB, alignment of the centerlines with the XA image is ensured. This enables direct comparison between the clinical images and the initial CoW model.

Next, algorithmic adjustments to the initial CoW model are made based on each XA image using 3D computer-aided design (CAD) software Rhino and Grasshopper as follows. The vessel diameters of the CoW model are measured at various locations along each vessel and projected onto the virtual detector plane of XA image. The vessel edges are manually drawn on the XA images and target vessel diameters are obtained at those locations. The model vessel diameters are then compared to the target vessel diameters to compute scale factors, which are used to locally adjust model vessel diameter radially about the vessel centerline.

Finally, to achieve a better agreement with XA images, iterative manual adjustments are made to the 3D model geometry. In this procedure, virtual angiographies of the 3D adjusted model are projected onto the virtual detector plane of each XA image using the alignments obtained earlier. The resulting 2D projections of the model are overlaid onto the XA images for comparison. In regions where the 2D projection does not match the vessel geometry as seen on the XA image, the model vessel geometry (i.e., vessel diameter and vessel curvature) is manually adjusted using MeshMixer. This adjustment-projection-comparison loop is iteratively repeated until the 2D projections of the model and the corresponding XA images agree, as confirmed by a neuroradiologist. The final adjusted model (**Supplementary Figure S1D)** is then exported as a triangulated surface mesh.

### 2.2 Reconstruction of solid NURBS mesh

The adjusted surface mesh is used to generate a volumetric nonuniform rational B-spline (NURBS) reconstruction of the CoW to be used in analysis. The methodology, detailed in a previous report^13^, uses an in-house template-based vascular modeling software that leverages the geometric modeling kernel of the CAD package Rhino for its robustness, interoperability, accuracy, and speed. First, centerlines are extracted from the triangulated surface mesh with a skeletonization algorithm. The centerlines are sampled at regular intervals, and at each sample point a perpendicular frame is constructed. At branch points, where multiple vessel centerlines meet, the frames are folded to build a conforming parameterization. A mesh-frame intersection curve is computed at each frame and interpolated with a NURBS curve. The NURBS curves are then lofted together along the centerlines to form a NURBS reconstruction of the triangulated surface mesh geometry that is parameterized in the circumferential and axial directions. Finally, a radial direction is added to the parameterization by extruding the surface control points to the centerline, yielding a hexahedral NURBS solid model (**Supplementary Figure S1E**) that is used for analysis. Mesh sizes and refinement strategies were chosen based on mesh independence studies (**Supplementary Figure S2**)^3^.

### 2.3 Governing equations and solution strategies

Blood flow was assumed to be governed by the unsteady Navier-Stokes equations with a time-dependent pulsatile inflow boundary condition^14^ imposed at the three inlets: left ICA (LICA), right ICA (RICA), and basilar artery (BA), a no-slip boundary condition was prescribed at the rigid vascular wall, and a traction-free outflow boundary condition implemented at the branch outlets (see **Fig. 1** for the simulation setup). Blood was modeled as an incompressible Newtonian fluid with a density *ρ* of 1060 kg/m^3^ and a dynamic viscosity *µ* of 0.0035 Pa×s. A Navier-Stokes solver within an isogeometric analysis framework^15–17^ was used to solve the system of equations by applying the solution strategies and numerical procedures described in a previous work^3^. The WSR was computed by using the following equation of wall shear stress vector ***τ*** = (***σ · n***) − ((***σ · n***) ***· n***)***n*** and the relation 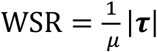, where (***σ · n***) is the traction vector, ***σ*** is the stress tensor, and ***n*** is the unit normal.

**Fig. 1:**
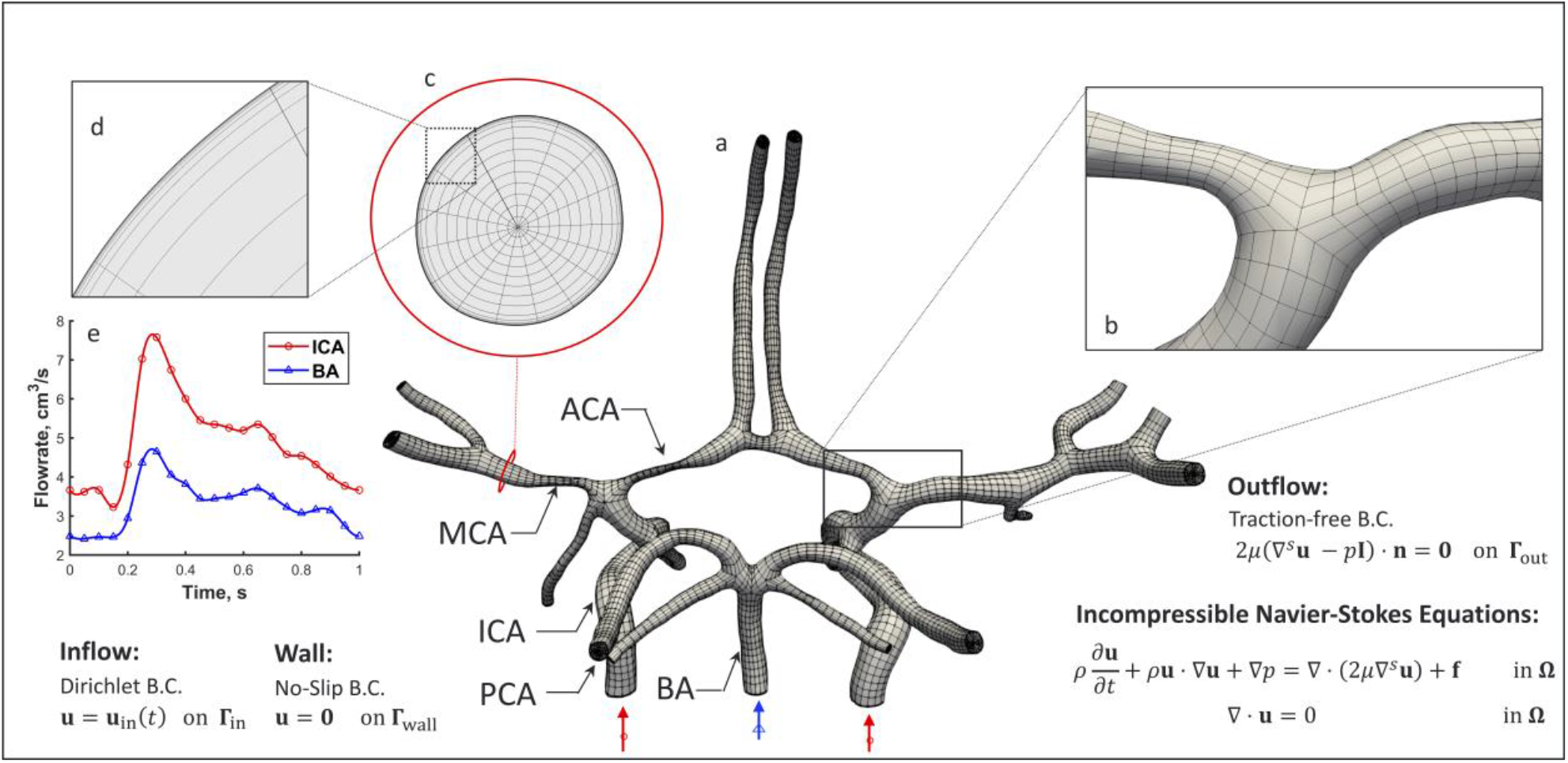
Simulation setup. (a) The hexahedral NURBS mesh of a human CoW with a bifurcation (b) is shown. A cross-section of the computational mesh (c) is shown to highlight the boundary layer refinement (d) we implemented. The governing equations subjected to boundary conditions (B.C.s) are also presented, where u represents velocity, *p* is the pressure, *f* is the external force, *μ* is dynamic viscosity, *ρ* is density, *t* is time, and n is the unit normal. (e) A pulsatile inflow condition is prescribed at the three inlets: left and right ICAs (red waveform) and BA (blue waveform). A no-slip B.C. is imposed on the vessel wall, and a traction-free B.C. is set at all the outlets.

## 3. Results

Data from 50 pediatric MMD patient were retrospectively reviewed (see **Supplementary Figure S3**)^3^ and 6 cases were selected for CFD analysis (**Table 1**) based on the following criteria: 1) patient age is less than 18 years, 2) no other cerebral or cerebrovascular disease is indicated, 3) history of at least one stroke, 4) availability of post-stroke MR TOF and XA imaging taken within 6 months of each other, and 5) there is communication between posterior and anterior circulations in the CoW. For each MMD patient, blood flow velocity fields were simulated in the CoW models reconstructed from imaging data and WSR values were quantified. The spatial distribution of WSRs at peak systole are shown in **Fig. 2**, along with timelines summarizing the chronology of stroke events, surgeries, and the collection of imaging data used to generate each model. The results are compared with those in a control subject (see **Supplementary Figure S4**).

**Table 1:** Summary of the MMD patient cohort (n = 6)

| ID | Gender | Primary Stroke | Suzuki Staging*** |  | Ipsilateral | Contralateral | Contralateral |
| --- | --- | --- | --- | --- | --- | --- | --- |
| | | Side | Ipsilateral | Contralateral | Surgery ($\Delta t^*$) | Stroke ($\Delta t^*$) | Surgery ($\Delta t^*$) |
| 1 | F | R | 2 | 2 | Yes (<1 yr) | Yes (<1 yr) | Yes (<1 yr) |
| 2 | F | R | 5 | 2 | Yes (<2 yr) | Yes<sup>a</sup> (<2 yr) | Yes (<3 yr) |
| 3 | M | L, L<sup>b</sup> | 2 | 0 | Yes (<1 yr) | Yes (<5 yr) | Yes (<3 yr) |
| 4 | F | R | 3 | 2 | Yes (<1 yr) | Yes<sup>c</sup> (<1 yr) | Yes (<1 yr) |
| 5 | M | L | 2 | 1 | Yes (<1 yr) | Yes<sup>d</sup> (<3 yr) | No |
| 6 | F | L | Atypical<sup>e</sup> |  | Yes (<1 yr) | No | Yes (<1 yr) |

\* $\Delta t$ is the time following primary stroke.
\*\* Suzuki staging performed by trained radiologists based on XA imaging used for model creation.
<sup>a</sup> Patient 2: Primary and secondary stroke prior to bilateral surgery and model image data collection.
<sup>b</sup> Patient 3: Two left side strokes within 6 months, prior to surgery. $\Delta t$ is based on first stroke in this case.
<sup>c</sup> Patient 4: Two contralateral, left side strokes.
<sup>d</sup> Patient 5: Two bilateral stroke events.
<sup>e</sup> Patient 6: Suzuki staging was not performed because of atypical anatomy.

**Fig. 2:**
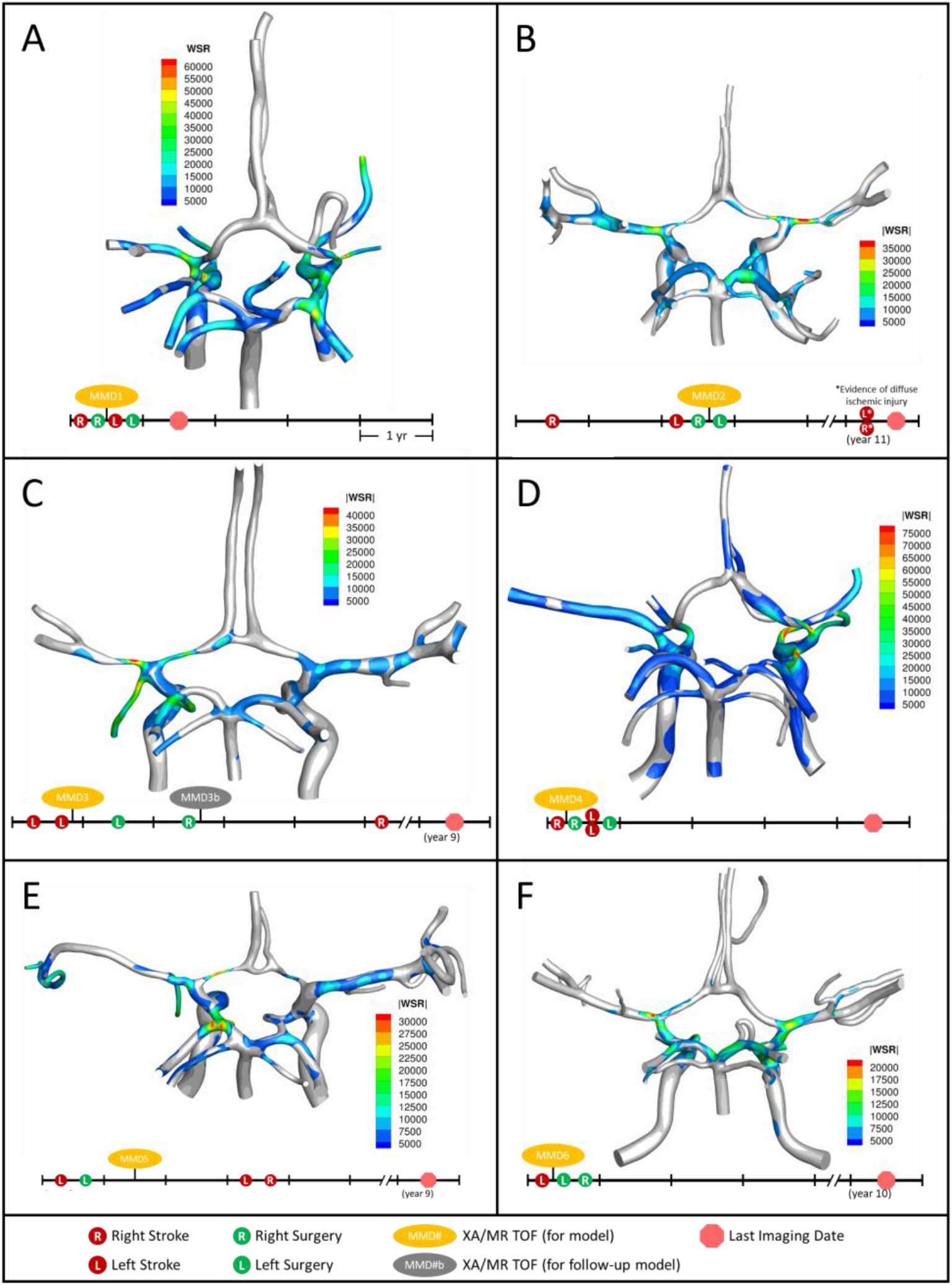
Distribution of WSR above the coagulation limit (> 5000 s^−1^) at peak systole (top) along with corresponding patient timeline (below) for the patients we studied: A) MMD1, B) MMD2, C) MMD3, D) MMD4, E) MMD5, F) MMD6. Red circles indicate stroke occurrence, green circles indicate surgery, yellow ovals denote model creation from XA and MR TOF imaging data, and red circle octagon is the last follow up imaging available.

Patient MMD1 presented with right-sided stroke and was treated with right dural inversion surgery. Post-operative images were used for CoW model creation and CFD analysis (**Fig. 2A**). At peak systole, simulation results predict extreme WSR levels in areas of vessel narrowing in right MCA (RMCA), right ACA (RACA) and RICA, (maximum 64,176 s^−1^, 61,294 s^−1^ and 44,617 s^−1^, respectively), coinciding with the location of the primary stroke. Concurrently, critical WSRs were seen in the terminal LICA (maximum 63,094 s^−1^). The patient experienced a contralateral stroke two months later.

Patient MMD2 initially presented with a right-sided stroke, followed by a left-sided stroke approximately two years later. A right dural inversion surgery was performed shortly after the second stroke. Post-operative images were used for model creation. At peak systole, WSR levels in the RACA and RMCA (**Fig. 2B**) reached 33,728 s^−1^ and 40,296 s^−1^, respectively, coinciding with areas with stenosis. Severe narrowing was also observed in the left ACA (LACA) (maximum WSR of 15,796 s^−1^).While not visibly impacted by MMD-related narrowing, the left MCA (LMCA) had elevated WSRs (maximum 28,370 s^−1^). A left dural inversion surgery was performed within one year of the left-sided stroke, and the disease stabilized during 7-year follow up, at the end of which, evidence of bilaterally diffuse ischemic injury was noted. It is unclear from the patient history if this is related to the previously mentioned bilateral stroke events.

Patient MMD3 suffered two left-sided strokes within the same year. Post-stroke images were used to create a CoW model. Extreme WSRs at peak systole were observed (**Fig. 2C**) in the LACA and LMCA (maximum 27,256 s^−1^ and 45,066 s^−1^, respectively), coinciding with the primary stroke location. Contralateral WSRs were considerably lower, peaking around 12,761 s^−1^ in the RMCA. Left-side and right-side dural inversion surgeries were performed within one and two years of the primary stroke, respectively. A right-sided stroke occurred approximately three years later.

Patient MMD4 presented with a right-sided stroke. Post-stroke images were utilized for model creation. Super critical WSR vales at peak systole were predicted (**Fig. 2D**) in the severely narrowed RACA (maximum 72,588 s^−1^) and in the RMCA (maximum 45,831 s^−1^) where an ischemic event was evident. Concurrently, elevated WSR levels were seen in the LMCA (21,203 s^−1^) and, despite showing vessel narrowing, the LACA showed no WSRs at peak systole above the coagulation threshold. Within a year of the primary stroke, patient underwent a right-side surgery, which was closely followed by two contralateral strokes. After a subsequent surgery on the left side, the disease stabilized.

Patient MMD5 suffered a left-sided stroke and within a year underwent left-side surgery. Post-operative images were utilized to create a CoW model. Elevated peak systolic WSR values (maximum 30,974 s^−1^) were predicted (**Fig. 2E**) in the severely narrowed region of the LACA and the LMCA (maximum 19,305 s^−1^). Contralaterally, the RMCA and the RACA, the latter of which showed significant stenosis, saw peak WSR values of 12,015 s^−1^ and 11,829 s^−1^, respectively. Within three years of the primary stroke, the patient suffered bilateral strokes.

Patient MMD6 presented with a left-sided stroke. Model created using post-stroke images shows that peak WSRs reached 22,140 s^−1^ in the LMCA, where there is a mild narrowing, and 18,060 s^−1^ in the terminal LICA (**Fig. 2F**). Contralaterally, the RMCA and terminal RICA had similarly elevated peak WSRs (17,865 s^−1^ and 20,980 s^−1^, respectively). Within one year of the primary stroke, bilateral dural inversion surgery was performed. The patient remained stable throughout the next 10 years of follow-up.

To further analyze the impact of elevated WSR values on stroke risk, we generate WSR distribution curves as described in **Figs. 3A** and **3B** for the right and left MCAs, ACAs, and terminal ICAs, where the terminal ICA is defined as the ICA segment between the ophthalmic artery and the ICA bifurcation. The WSR distribution curve for a vessel shows how much of the vessel’s surface area (SA) exceeds a given WSR threshold value. **Figs. 3C** and **3D** illustrates these curves for the left side vessels of the control subject and patient MMD3, respectively, indicating that vessel areas of higher WSR are more widespread for patient MMD3 compared to the control. From the WSR distribution curves, two metrics are derived to assess stroke risk.

**Fig. 3:**
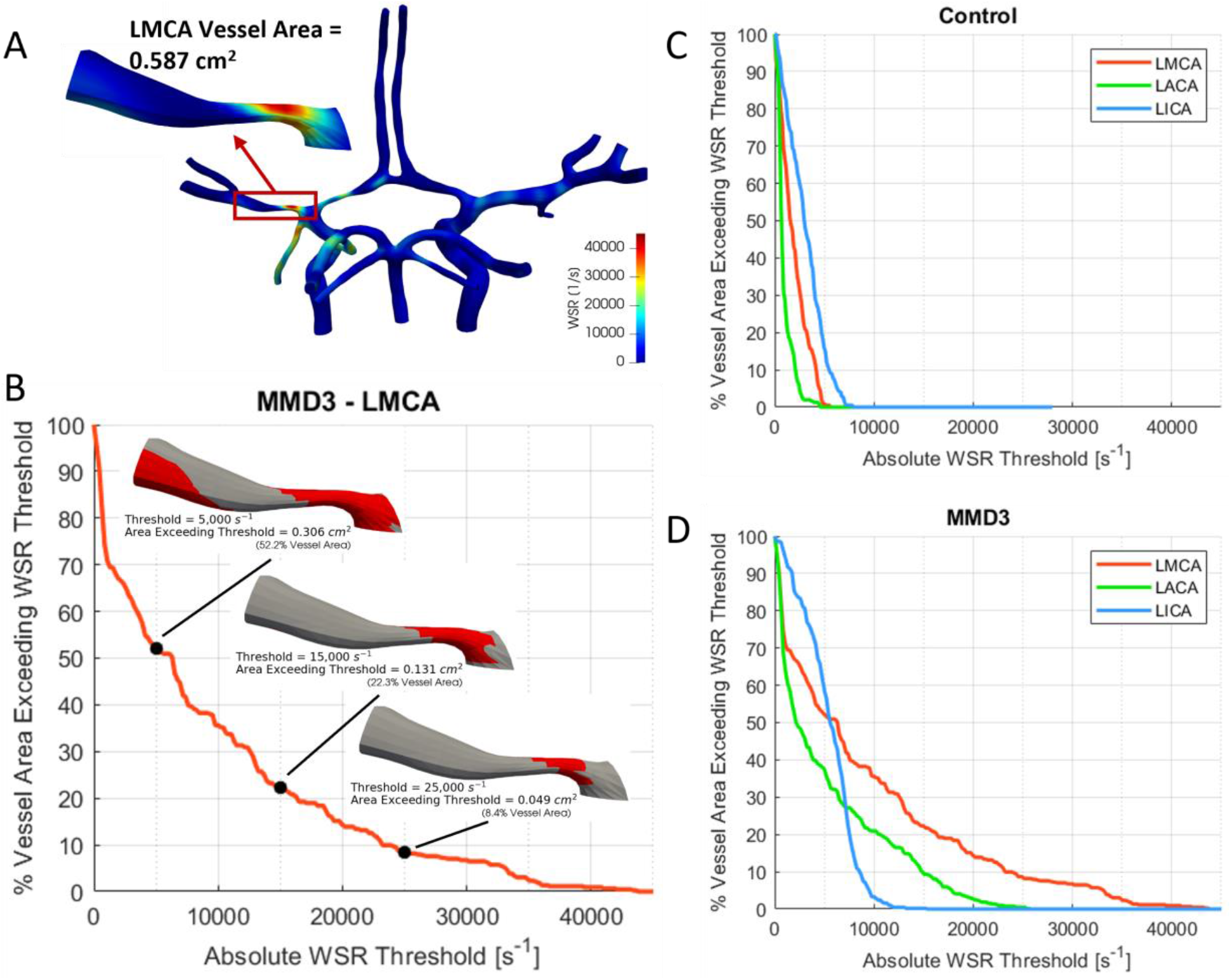
a) Construction of WSR curve. First, a vessel segment is extracted from the model and the total surface area is measured. A WSR threshold is set and the surface area that exceeds this threshold is quantified and then divided by the vessel’s total surface area to obtain percent vessel area exceeding WSR threshold, which is then calculated for multiple threshold values ranging from 0 to 100,000 s^−1^ with an increment as low as 100 s^−1^ to generate the WSR curve. The WSR curves for the left and right ICAs, MCAs and ACAs are shown for c) the control subject and d) MMD3.

First, we consider the “critical WSR coverage” (CWSRC) in each vessel representing the percent of each vessel’s surface area with critical WSR values – i.e., those exceeding the coagulation threshold of 5000 s^−1^ (**Fig. 4)**. A graphical explanation of how this metric is derived from the WSR distribution curve is shown in **Supplementary Figure S5**. In the control, three vessels (LMCA, LACA, and RACA) had little to no surface area with critical WSRs (0.9%, 0.0%, and 0.0%, respectively). The remaining vessels under consideration have mild to moderate CWSRC (LICA: 15.8%, RMCA: 29.0%, RICA: 35.9%). In contrast, many of the vessels in the 6 MMD patients exhibit greater CWSRC than those in the control. Of note, the RMCA of MMD1 saw critical WSRs covering 88.3% of its surface area, and MMD4 saw high CWSRC in the LMCA (77.1%) and the RMCA (96.5%). As an aggregate form of this metric, the three vessels on either side are combined to form two regions, left and right, in which the CWSRC is assessed (**Supplementary Figure S6**). For the control, the aggregate CWSRC values were 6.3% and 20.5%, for the left and right sides, respectively. In the MMD patients, these aggregates vary from 22% to 52.2% on the left side and 13.7% to 63.7% on the right side, and each patient has at least one side with an aggregate above 20.5%, the greater of the two aggregates in the control.

**Fig. 4:**
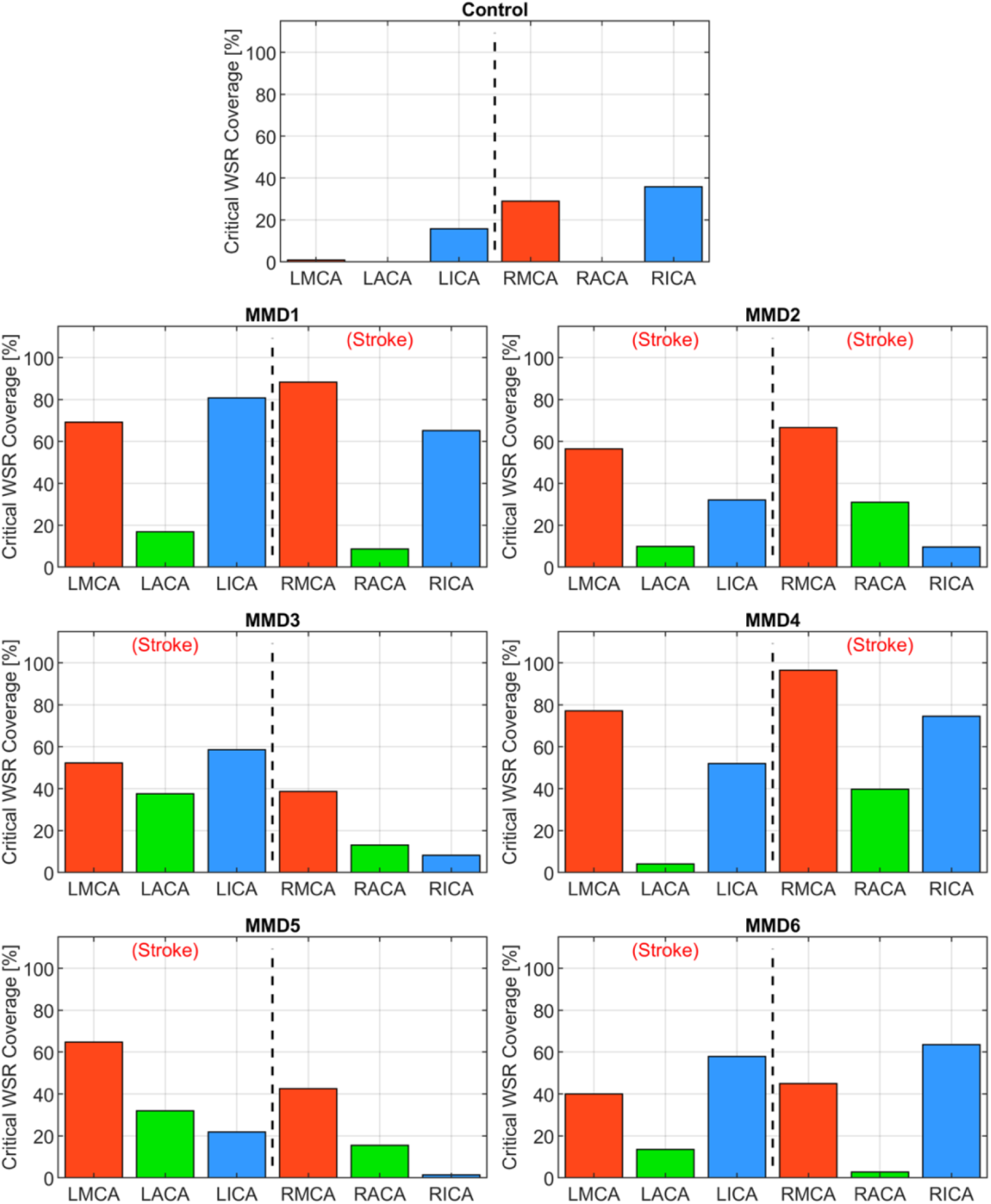
Critical WSR coverage, expressed as a percentage of each vessel’s surface area with WSR 5000 s^−1^, in the left and right MCAs, ACAs, and ICAs for the control subject and the 6 MMD patients we studied. The label (Stroke) indicates the side of primary stroke.

The second metric we examine is a “WSR score” for each vessel calculated by integrating the WSR distribution curve over the domain [5000, ∞) above the coagulation limit (see **Supplementary Figure S7**). This parameter gives an indication of the extent and magnitude of the higher WSRs. For example, two vessels with the same CWSRC can have different WSR scores if one of them has higher WSR values occupying the same vessel surface area. WSR score is relatively small or zero for the vessels in the control while many of the vessels in the MMD patients have significantly higher WSR scores with each having at least two vessels with a WSR score at least 4x the maximum seen in the control, and all but one (MMD6) with at least one vessel with a WSR score greater than 8x the maximum control WSR score **(Fig. 5)**. The MMD patients have left and right side aggregate WSR scores ranging from 1.5x to 37.5x the control’s largest aggregate WSR score **(Supplementary Figure S8)**.

**Fig. 5:**
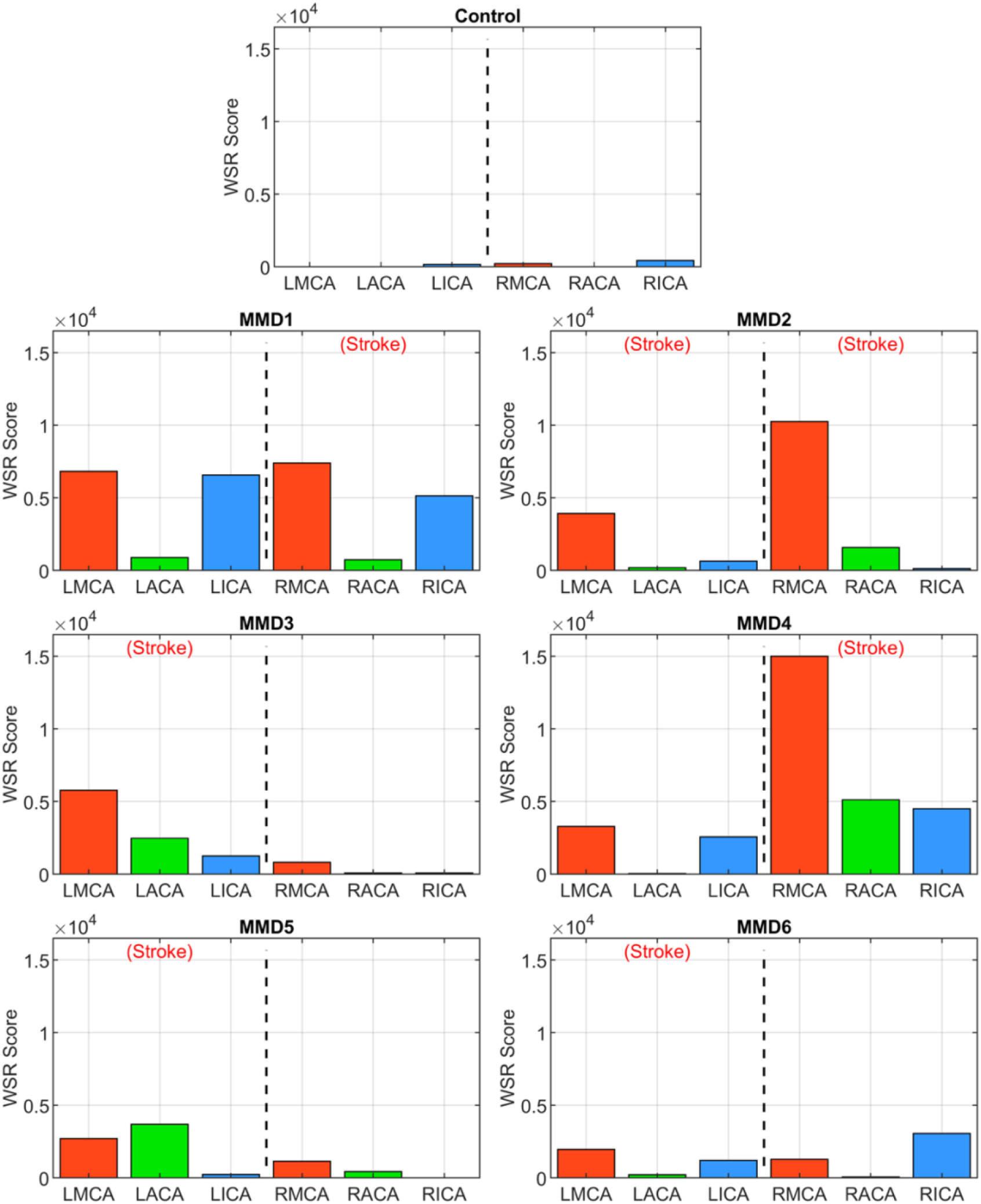
WSR score in the left and right ICAs, MCAs and ACAs for the control subject and the 6 MMD patients we studied. The label (Stroke) indicates the side of primary stroke.

**Fig. 6** summarizes a longitudinal study conducted for patient MMD3 where a second CoW model (MMD3b) was generated using imaging data collected soon after the 2^nd^ (right-side) dural inversion surgery to assess how disease progression impacts the WSR-based metrics for predicting future stroke risk. The most notable change is that the LACA has become fully occluded. Additionally, corresponding with approximately two years of patient growth, the vessels of the second model generally have larger diameters. As a result, the WSR values predicted at peak systole are less extreme for MMD3b (global maximum WSR of approximately 17,000 s^−1^) compared to MMD3 (45,000 s^−1^). On the left-side, marked reductions in both metrics were seen compared to the initial model MMD3 (**Figs. 6H** and **6I**). CWSRC decreased in the LMCA (down to 12.9% from 52.2%) and in the terminal LICA (29.1% versus 58.5%), and the left-side aggregate CWSRC decreased from 52.1% to 22.0%. The WSR scores in the LMCA and LICA also decreased by 87% and 71%, respectively, and the aggregate WSR score on the left decreased by approximately 88%. Contralaterally, a larger surface area of the RMCA and RACA exceeded the coagulation threshold (from 38.7% to 46.8% and from 13.1% to 28.9%, respectively), and the aggregated CWSRC on the right increased from 17.6% to 18.7%. The WSR score in the RMCA and RACA were approximately 3x and 17x that of the first model, respectively, and the right-side aggregate WSR score was approximately 3.7x that of the first model.

**Fig. 6:**
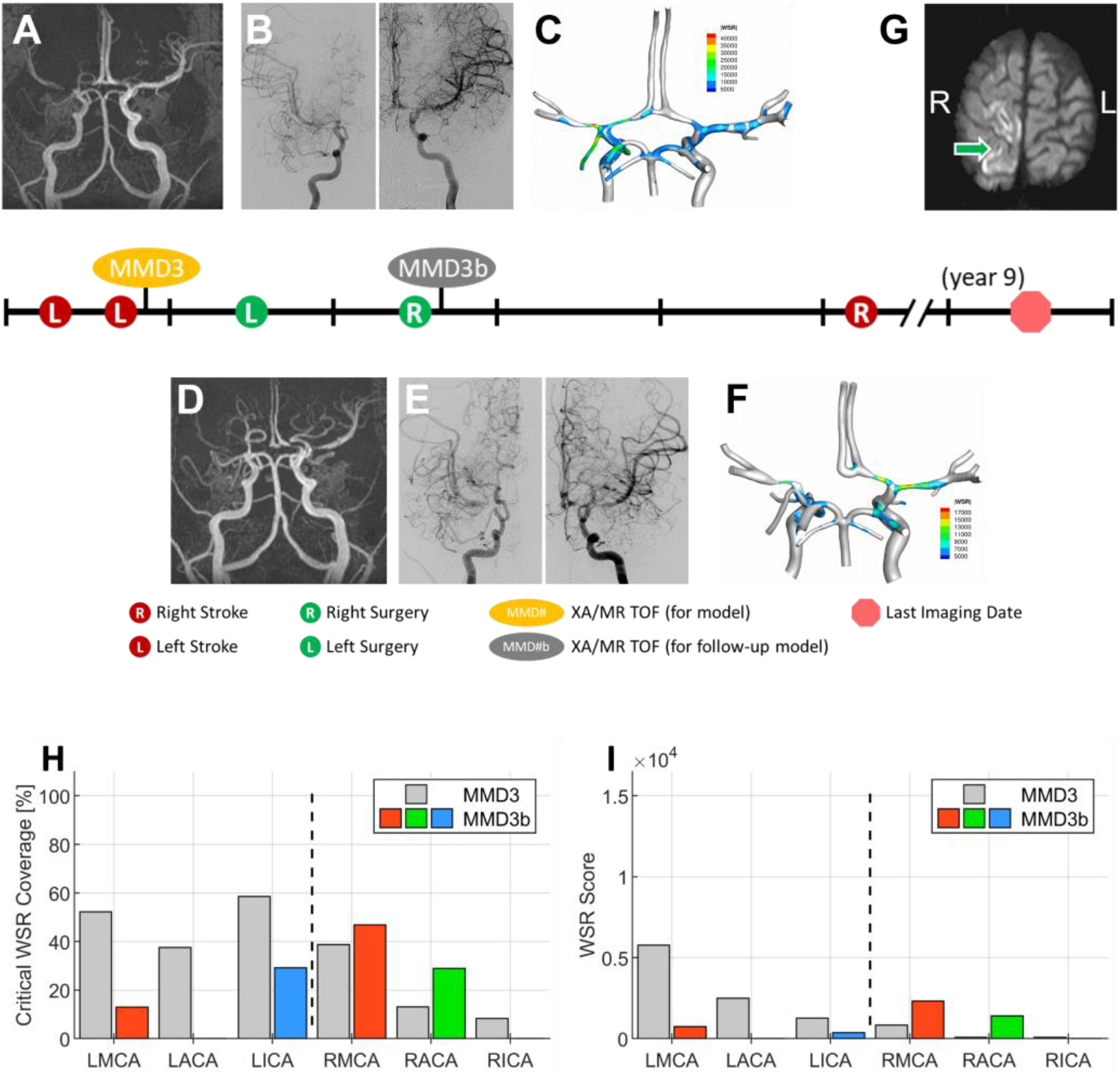
A representative case. Patient presented with multiple transient ischemia attacks on the left side. Initial brain MR TOF (A) and XA (B) confirmed occlusion of left M1 and left A1, and mild to moderate narrowing of right A1 and right M1 segments. The patient underwent unilateral left dural inversion. (C) WSR distribution pattern in the patient’s CoW model (MMD3) created from postoperative images. The follow-up MR TOF (D) and XA (E) imaging within two years from the initial diagnosis show evidence of moderate contralateral progression of disease and right dural inversion surgery. (F) WSR distribution pattern in the CoW model (MMD3b) created from postoperative images. In the follow-up transverse (axial) soft tissue MR image (SENSE sequence) at approximately 5 years from the initial diagnosis, evidence of right-sided stroke within perirolandic region involving right MCA territories (G). Plots showing the critical WSR coverage (H) and WSR scores (I) of each vessel for the second CoW model (MMD3b) with comparison to the results from MMD3 (gray bars).

## 4. Discussion

We performed blood flow analysis of patient-specific models of the CoW in pediatric MMD patients and showed that local hemodynamics are markedly different than those present in a healthy subject^3^. These differences, which are largely dependent on vessel morphology altered by MMD progression, may be indicative of future contralateral stroke risk in patients presenting with unilateral stroke. We hypothesized that a critically high WSR above the coagulation limit of 5000 s^−1^ could cause thrombus formation, potentially leading to an ischemic event^8^ and a careful analysis of local WSR could be predictive of future stroke. As such, we focused on the spatial distribution of WSR of the major vessels of the anterior circulation: ICA, MCA, and ACA^18^ and considered two WSR-derived metrics. First, we computed CWSRC, the percentage of the vessel’s surface area with critical WSR values exceeding the coagulation threshold. Second, we calculated a WSR score that gives a sense of the magnitude and extent of the critically high WSR values present in a vessel.

Among the 6 patients we selected for analysis, all of which had at least one unilateral stroke event prior to the collection of the images used for model creation, three (MMD1, MMD3, and MMD4) suffered contralateral stroke after being surgically treated on the affected side, one (MMD5) had bilateral stroke after being surgically treated on the affected side, one (MMD2) experienced bilateral stroke prior to the imaging used for model creation, and one (MMD6) had no additional strokes within the ten years of follow-up after being treated with bilateral surgery. In the patient with a history of bilateral stroke (MMD2), both right and left MCAs have more than 50% CWSRC and WSR scores > 9x the max WSR score of the vessels in the control. The metrics are similarly high in the other patients in the vessels ipsilateral to the primary stroke. One or more vessels with 50% surface area with critical WSRs and WSR scores > 7x the maximum control WSR score are seen in all these cases, which is consistent with the stroke events the patients experienced.

In two patients (MMD1 and MMD4), contralateral strokes occurred shortly after the primary stroke and being surgically treated on the affected side. High metric values (more than 50% CWSRC and WSR scores greater than 6x the max control WSR score) were seen in at least two of the contralateral vessels, potentially indicative of the impending contralateral stroke events. As further evidence of contralateral stroke risk for MMD1, the aggregated CWSRC on the contralateral side (52.2%) is greater than that on the primary stroke side (41.4%), and the aggregate WSR score, which is approximately 22x the maximum aggregated WSR score in the control, is also 7.5% higher on the contralateral side. This may indicate that at the timepoint represented by this CoW model, the disease state on the left side may have become more severe than what caused the primary right-sided stroke. On the other hand, for MMD4, the aggregate CWSRC on the contralateral side is comparable to the ipsilateral side (50.6% vs. 63.7%, respectively), but the contralateral aggregate WSR score is only 24% of the ipsilateral side. However, this value is still approximately 9x that seen in the control, indicating that increased contralateral stroke risk may exist.

For the other two patients (MMD3 and MMD5) that experienced contralateral strokes, these metrics do not strongly suggest future contralateral stroke. There were no contralateral vessels with >50% CWSRC or with WSR scores >3x the max control WSR score. The contralateral aggregate CWSRCs are much less when compared to the stroke-side for MMD3 (17.6% vs. 52.1%) and MMD5 (13.7% vs. 29.1%) and the aggregate WSR scores in vessels contralateral to the primary stroke side are 10% and 24% of the ipsilateral aggregates for MMD3 and MMD5, respectively. While none of the WSR metrics strongly indicate future stroke risk, in both cases, there is a multi-year gap between the imaging used for model creation and the subsequent contralateral stroke(s). This suggests that perhaps we can reasonably predict future strokes in the near term, and that additional modeling, at additional timepoints during follow-up, may be warranted for patients without conclusively high or comparable (to ipsilateral) metric values.

To assess how local hemodynamics changes over time as the disease progresses, we examined imaging data at a subsequent time point for patient MMD3, approximately two years after the primary stroke. Comparing the two modeling timepoints (i.e., MMD3 and MMD3b), both CWSRC and WSR score decrease for all three vessels ipsilateral to the primary stroke, possibly indicating that the disease is stabilizing on the left side. Conversely, marked increases in the two metrics are seen in the RMCA and RACA. The aggregate WSR score on the contralateral side is > 3x the score on the left side, suggesting that the patient is at risk of a contralateral stroke. Patient MMD3 did indeed go on to have a right-sided stroke three years later. Overall, the results from this longitudinal study of patient MMD3 demonstrate the utility in regular patient monitoring and performing hemodynamic analysis using updated CoW models to assess changes in stroke risk over time.

Assessing multiple timepoints may have been particularly useful for patient MMD6. The CWSRC in the vessels contralateral to the primary stroke were comparable to the ipsilateral side (35.0% vs. 39.8%, respectively). While the same pattern holds for the WSR scores, aggregate WSR score is approximately 31% greater on the contralateral side. Although similar evidence in other patients were predictive of future contralateral stroke, no additional stoke events are noted in this patient’s history. Notably, this patient had a bilateral surgery shortly after the primary stroke, which is known to improve cerebral perfusion and help washout thromboemboli in MMD patients^8^, thereby potentially preventing further stroke.

In this work, we adopt a WSR of 5000 s^−1^ as a limit for coagulation, above which localized platelet accumulation can occur on a thrombogenic surface^7^. However, in the MMD patients we studied, stroke events did not seem to occur immediately upon exceeding this coagulation limit. The lowest WSR that could be associated with a subsequent contralateral stroke was approximately 21,000 s^−1^ (MMD4). In patient MMD1, even though the contralateral WSRs reached 12x the coagulation limit and 5x that seen in the control subject, contralateral stroke did not occur until two months later. The mechanisms of stroke in MMD are still unclear^8^. Microfluidics models studying the effect of WSR on platelet rich thrombus formation suggest that pathologically high flow rates induce endothelial damage, platelet aggregation, and fibrin disposition^19^. In MMD, pathogenesis of stroke could be explained as follows. Upon thrombus formation, entrapped cells remain in the intimal layer even after the clot resolves. With repeated clot formation, intimal hyperplasia results and the lumen narrows. An acute stroke can result when a clot or embolus finally occludes the lumen. Further studies of MMD pathology are required to understand this phenomenon.

The study presented here analyzed a limited cohort selected from a single-hospital dataset, and thus, our findings are likely limited by selection bias and the inherent biases associated with a retrospective study. Future studies are needed with larger sample sizes to confirm the correlations identified here between WSR and MMD progression and future stroke risk. This should include analysis of how timing of neurosurgical intervention impacts modeling results. In this retrospective study, due to limited availability of imaging data, the models were created irrespective of when surgery occurred, if it occurred at all. Future prospective studies should also aim to measure flowrates in the ICAs and BA at time of imaging (e.g., including 4D MRI sequences to the MR TOF acquisition) so that patient-specific inlet velocities can be prescribed in the simulations.

In summary, we describe two WSR-based metrics for assessing stroke risk in pediatric MMD patients. In a subset of the patients evaluated, these metrics strongly predict imminent contralateral stroke as confirmed by patient outcome data. Inconclusive results in the remaining patients highlight the difficulty in making predictions based on the hemodynamic environment at a single time point. Changes in the WSR-based metrics over time could provide a more accurate assessment of future stroke risk. Given the limited number of patients studied herein, a prospective longitudinal study of a larger randomized patient cohort is warranted. If shown promise in the larger study, our patient-specific computational modeling approach could be used to noninvasively predict future stroke occurrences in pediatric MMD patients and help ensure that unilateral MMD patients are properly monitored and treated before strokes occur.

## Supporting information

Electronic Supplemental Material

## Data Availability

All data produced in the present study are available upon reasonable request to the authors

## Abbreviations

MMD: Moyamoya Disease
CoW: Circle of Willis
WSR: Wall Shear Rate
ICA: Internal Carotid Artery
MCA: Middle Cerebral Artery
ACA: Anterior Cerebral Artery
BA: Basilar Artery
CAD: Computer Aided Design
NURBS: Non-Uniform Rational B-Splines
CWSRC: Critical Wall Shear Rate Coverage

## Acknowledgements

The authors gratefully acknowledge Karen Chen, MD at Texas Children’s Hospital for helping with Suzuki staging, Mario Dada at the University of Texas at Austin for assisting with image segmentation and Texas Advanced Computing Center (TACC) for providing high-performance computing resources.

## Ethical approval

There was no direct involvement of human subjects or protected health information (PHI). All patient imaging data used in analysis and modeling was collected retrospectively from medical records and de-identified at Texas Children’s Hospital. Institutional review board (IRB) approval was obtained with a waiver of written authorization for consent.

## Supplemental Material

Supplemental material for this article is available online.

## REFERENCES

1. Hirotsune N, Meguro T, Kawada S, Nakashima H, Ohmoto T. Long-term follow-up study of patients with unilateral moyamoya disease. Clinical neurology and neurosurgery. 1997;99:S178–S181

2. Hayashi K, Suyama K, Nagata I. Clinical features of unilateral moyamoya disease. Neurologia medico-chirurgica. 2010;50:378–385

3. Hossain SS, Starosolski Z, Sanders T, Johnson MJ, Wu MCH, Hsu M-C, et al. Image-based patient-specific flow simulations are consistent with stroke in pediatric cerebrovascular disease. Biomechanics and Modeling in Mechanobiology. 2021

4. Zipfel GJ, Sagar J, Miller JP, Videen TO, Grubb RL, Jr., Dacey RG, Jr., et al. Cerebral hemodynamics as a predictor of stroke in adult patients with moyamoya disease: A prospective observational study. Neurosurg Focus. 2009;26:E6

5. Yeon JY, Shin HJ, Kong DS, Seol HJ, Kim JS, Hong SC, et al. The prediction of contralateral progression in children and adolescents with unilateral moyamoya disease. Stroke. 2011;42:2973–2976

6. Derdeyn CP. Hemodynamic impairment and stroke risk: Prove it. AJNR Am J Neuroradiol. 2001;22:233–234

7. Casa LD, Deaton DH, Ku DN. Role of high shear rate in thrombosis. J Vasc Surg. 2015;61:1068–1080

8. Kim DY, Son JP, Yeon JY, Kim G-M, Kim J-S, Hong S-C, et al. Infarct pattern and collateral status in adult moyamoya disease. Stroke. 2017;48:111–116

9. Milewicz DM, Ostergaard JR, Ala-Kokko LM, Khan N, Grange DK, Mendoza-Londono R, et al. De novo acta2 mutation causes a novel syndrome of multisystemic smooth muscle dysfunction. Am J Med Genet A. 2010;152A:2437–2443

10. Starosolski Z, Villamizar CA, Rendon D, Paldino MJ, Milewicz DM, Ghaghada KB, et al. Ultra high-resolution in vivo computed tomography imaging of mouse cerebrovasculature using a long circulating blood pool contrast agent. Sci Rep. 2015;5:10178

11. Lee WJ, Jeong SK, Han KS, Lee SH, Ryu YJ, Sohn CH, et al. Impact of endothelial shear stress on the bilateral progression of unilateral moyamoya disease. Stroke. 2020;51:775–783

12. Hoogeveen RM, Bakker CJG, Viergever MA. Limits to the accuracy of vessel diameter measurement in mr angiography. Journal of Magnetic Resonance Imaging. 1998;8:1228–1235

13. Urick BY, Sanders TJ, Hossain S, Zhang Y, Hughes TJ. Review of patient-specific vascular modeling: Template-based isogeometric framework and the case for cad. Archives of Computational Methods in Engineering. 2019;26:381–404

14. Wahlin A, Ambarki K, Birgander R, Wieben O, Johnson KM, Malm J, et al. Measuring pulsatile flow in cerebral arteries using 4d phase-contrast mr imaging. AJNR Am J Neuroradiol. 2013;34:1740–1745

15. Hossain SS, Hossainy SFA, Bazilevs Y, Calo VM, Hughes TJR. Mathematical modeling of coupled drug and drug-encapsulated nanoparticle transport in patient-specific coronary artery walls. Computational Mechanics. 2012;49:213–242

16. Hossain SS, Zhang Y, Liang X, Hussain F, Ferrari M, Hughes TJ, et al. In silico vascular modeling for personalized nanoparticle delivery. Nanomedicine (Lond). 2013;8:343–357

17. Hsu M-C, Akkerman I, Bazilevs Y. High-performance computing of wind turbine aerodynamics using isogeometric analysis. Computers and Fluids. 2011;49:93–100

18. Kelly ME, Bell-Stephens TE, Marks MP, Do HM, Steinberg GK. Progression of unilateral moyamoya disease: A clinical series. Cerebrovascular diseases. 2006;22:109–115

19. Tsai M, Kita A, Leach J, Rounsevell R, Huang JN, Moake J, et al. In vitro modeling of the microvascular occlusion and thrombosis that occur in hematologic diseases using microfluidic technology. J Clin Invest. 2012;122:408–418

