## Supplementary material for "Toward noninvasive assessment of stroke risk in pediatric cerebrovascular disease": Electronic Supplemental Material

ELECTRONIC SUPPLEMENTARY MATERIAL

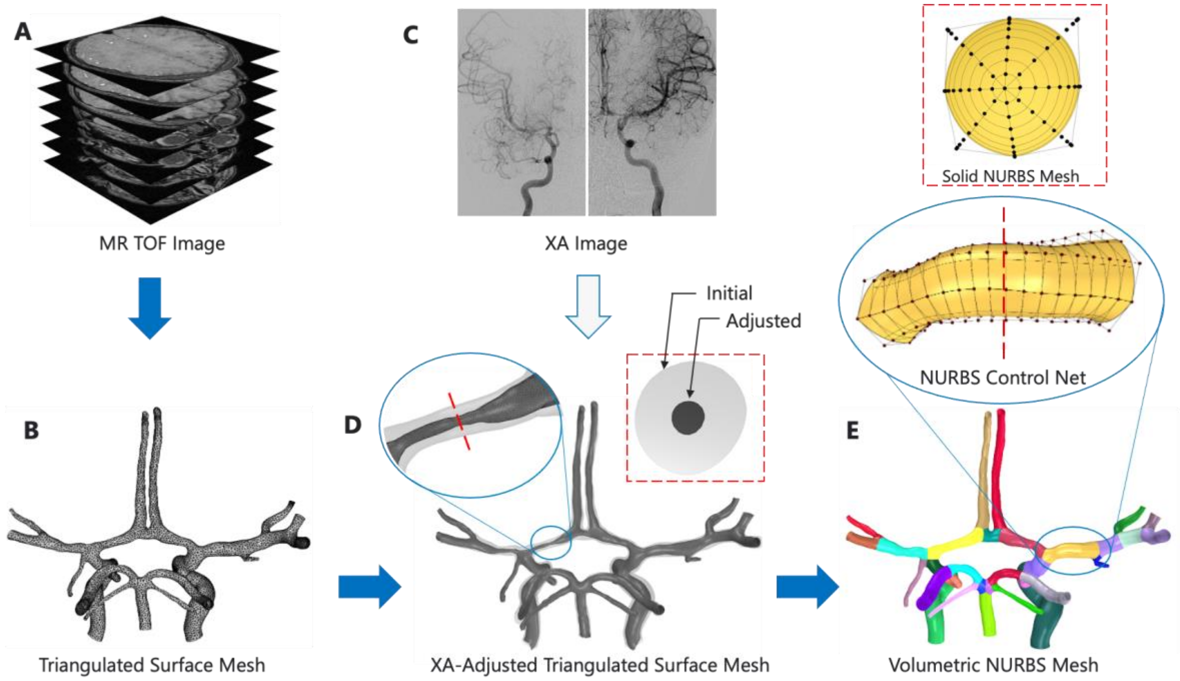


**Supplementary Figure S1:** Creation of circle of Willis (CoW) model from imaging data. The CoW vasculature is segmented from an MR time of flight (TOF) image stack (A) and exported as a triangulated surface mesh (B) using 3D Slicer. The segmented mesh is then registered to (C) X-ray angiography (XA) images by aligning its corresponding 2D projections. The diameters along the mesh are locally adjusted to match the diameters of the XA images. (D) shows the adjustments made across the entire mesh. Inset highlights the adjustments made along the left anterior cerebral artery (ACA) along with a cross section of left ACA where the diameter required a 70% adjustment. Centerlines are extracted from the adjusted surface mesh and used to construct a Nonuniform Rational B-Splines (NURBS) reparameterization (E) of the adjusted triangulated surface mesh. Inset shows a single NURBS patch and its control net along with a cross-section. To construct a volumetric mesh to be used in a computational fluid dynamics analysis, the control points of each axial frame in the NURBS surface control net are extruded in the radial direction to a point on the centerline.

A

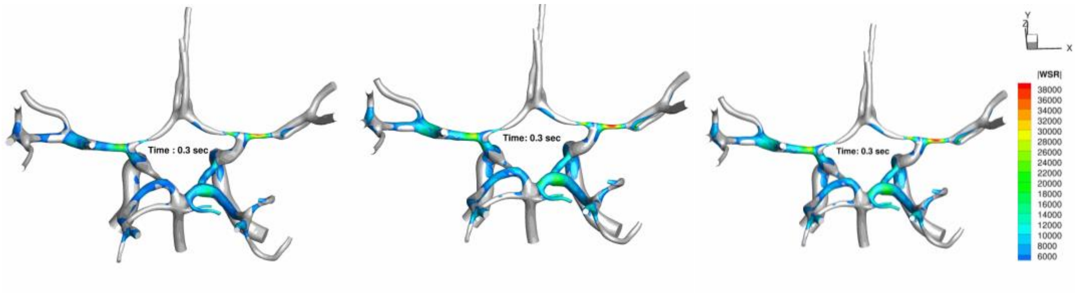


B

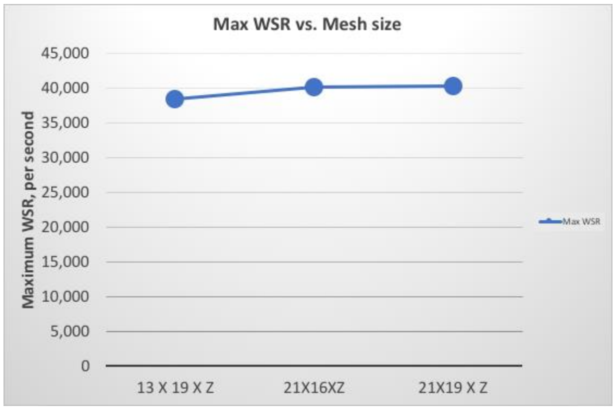


**Supplementary Figure S2:** Mesh independence study. (*a*) Wall shear rate (WSR) distribution in the CoW model of patient MMD3 for Refinement Mesh 1 (13 × 13 × Z), Refinement Mesh 2 (21 × 16 × Z), and Refinement Mesh 3 (21 × 19 × Z), from left to right. (*b*) Maximum WSR value in the right middle cerebral artery at peak systole for Refinement Meshes 1, 2, and 3 (left to right).

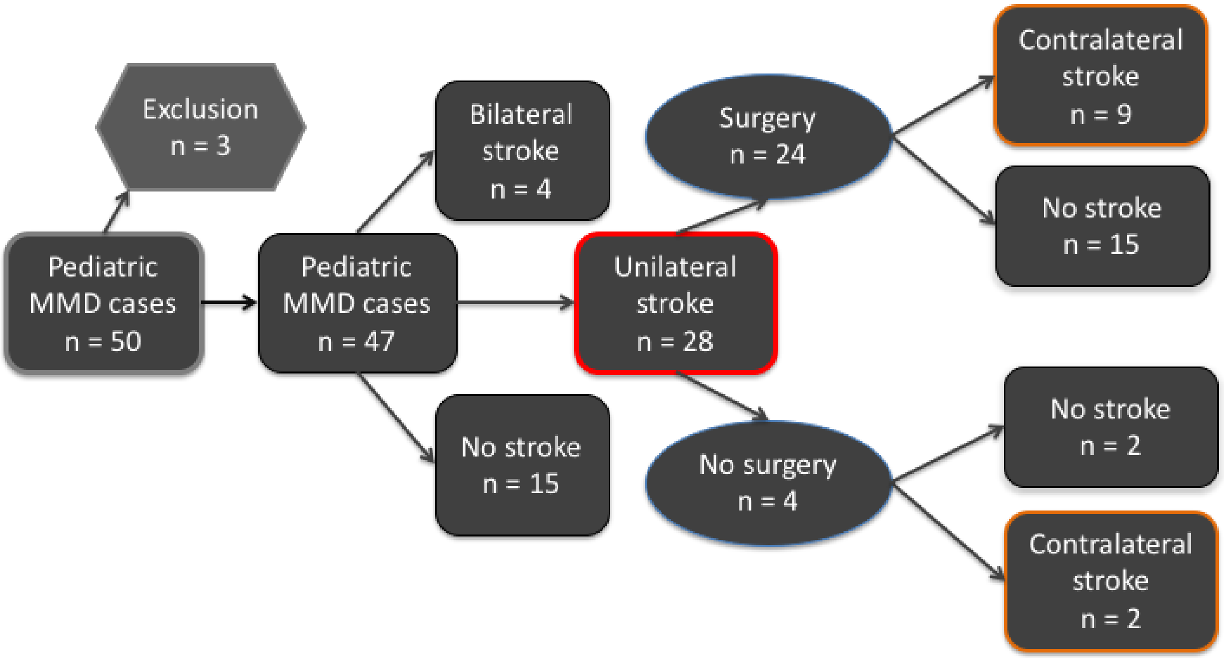


**Supplementary Figure S3:** A retrospective study of 50 pediatric MMD cases with known stroke history who were followed up longitudinally 2.79±2.15 years on average. Forty-four percent of these patients were female (mean age 9.82±-5.63 years), while 56% were male (mean age 10.30±5.0 years). Three cases were eliminated due to lack of adequate patient data. Among the remaining patients (n = 47), 15 (30.9%) did not have a stroke, 4 (8.5%) had bilateral stroke (stroke on both sides) and 28 (60%) suffered a unilateral stroke. 24 (85.7%) of the unilateral stroke patients underwent post-stroke dural inversion surgery. However, 11 (39.2%) of these unilateral stroke patients, 9 of whom underwent dural inversion surgery, suffered a subsequent contralateral stroke. Three (27%) of the contralateral stroke patients underwent another revascularization surgery. Among the remaining 8 (73%) contralateral stroke patients who did not undergo a second post-stroke surgery, two (25%) suffered recurrent strokes, either contralateral, or ipsilateral. These observations suggest the possibility that treatment by dural inversion can be followed by a contralateral stroke.

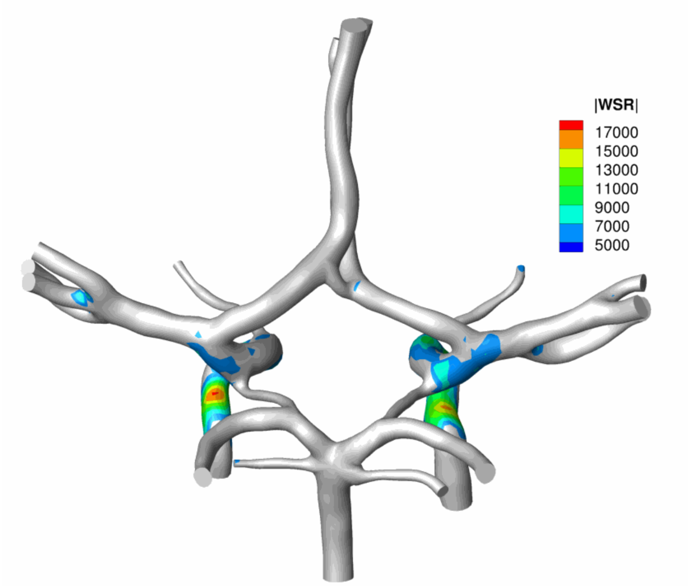


**Supplementary Figure S4:** Critical WSR (> 5000 $s^{-1}$) distribution at peak systole ($t$ = 0.3 s) in the CoW of a control subject.. While a global maximum WSR of approximately 17,000 $s^{-1}$ was predicted, the WSR does not exceed 11,000 $s^{-1}$ in the vessels under consideration : terminal internal carotid artery (ICA), proximal middle cerebral artery (MCA), and proximal ACA.

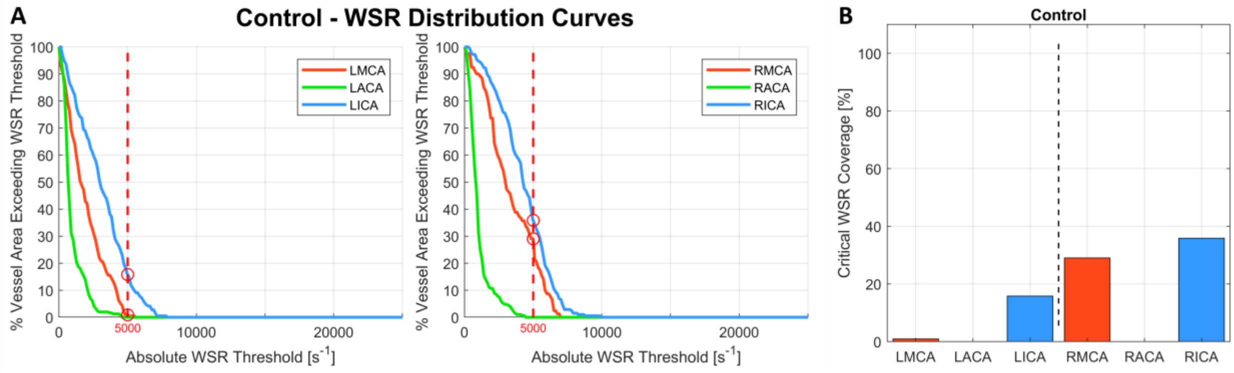


**Supplementary Figure S5:** Computation of the critical WSR coverage for the vessels of the control subject. This metric is percentage of a vessels surface area with WSR values exceeding the coagulation threshold (5000 s$^{-1}$) and evaluation the WSR distribution curve for each vessel (A) at a WSR threshold of 5000 s$^{-1}$ yields the critical WSR coverage values shown in (B) for the vessels of the control model.

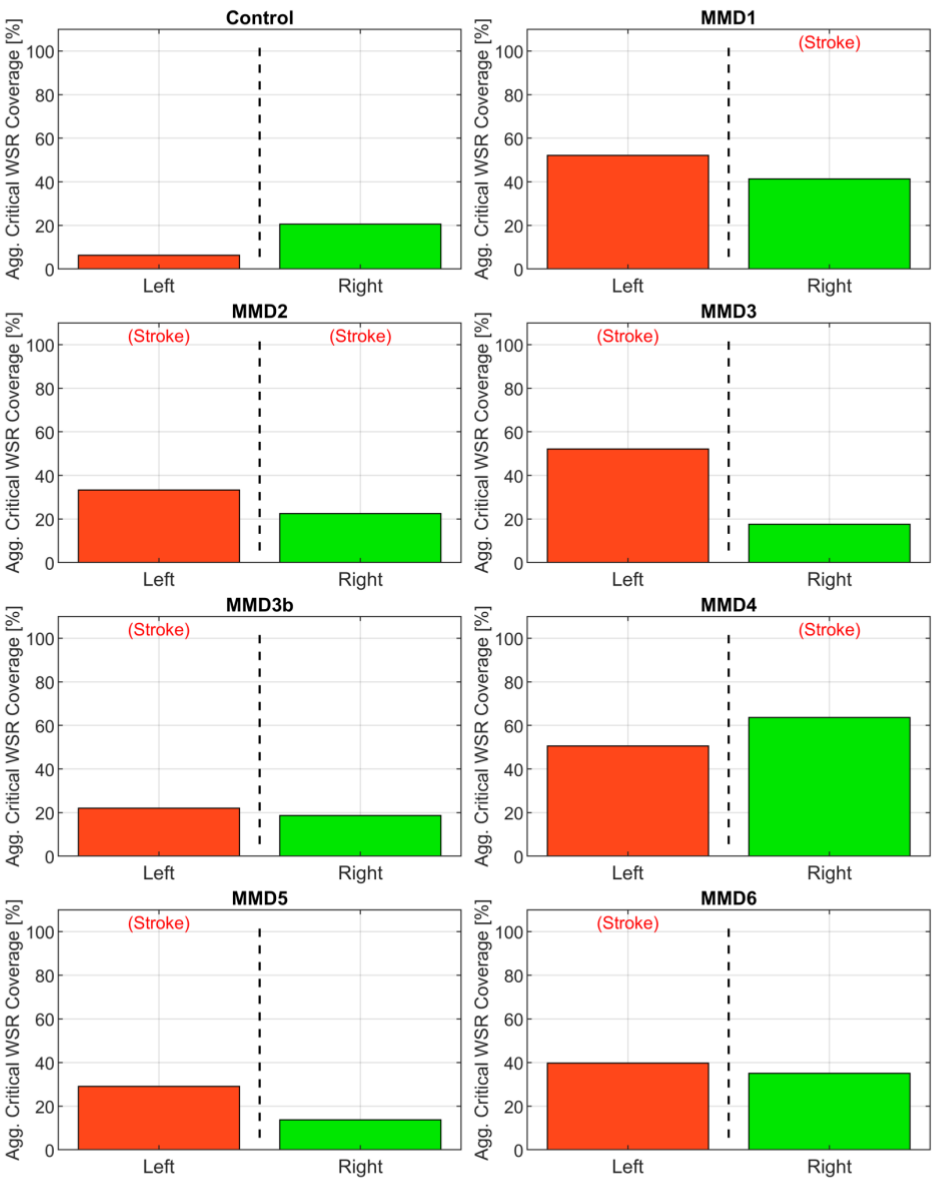


**Supplementary Figure S6:** Aggregate critical WSR coverages for the left and right sides of the anterior circulation (MCA, ACA, and ICA) for the control subject and the 6 patients we studied: MMD1, MMD2, MMD3 and MMD3b, MMD4, MMD5 and MMD6. The percentage is calculated as the sum of the surface area exceeding WSR > 5000 $s^{-1}$ in the three vessels divided by the sum of the surface areas of the three vessels. The label (Stroke) indicates the side of primary stroke event.

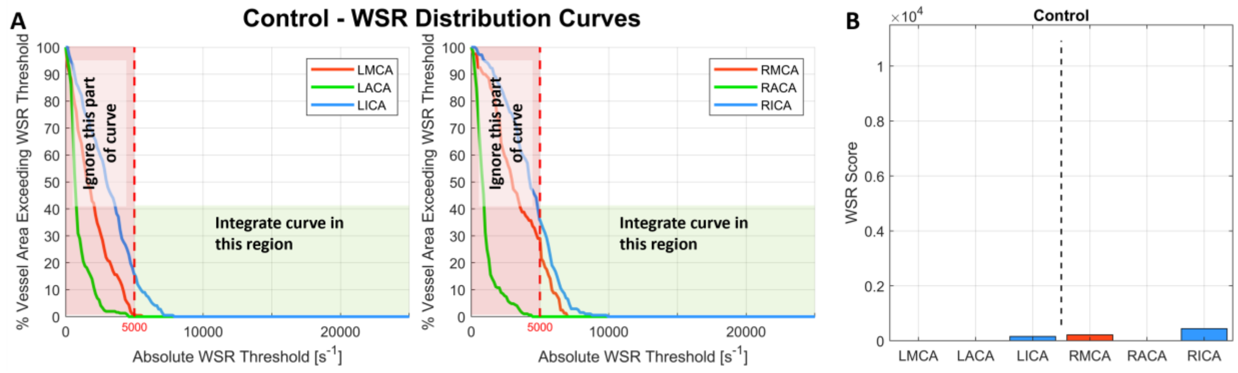


**Supplementary Figure S7:** Wall shear rate (WSR) score computation. (A) The WSR distribution curves generated for each vessel are integrated in the domain where WSR threshold is above 5000 $s^{-1}$ (the green area under the curve). The portion of the curve with WSR threshold value less than 5000 $s^{-1}$ (in red) is ignored. (b) WSR score for the left and right ICA, MCA and ACAs of the control subject are shown.

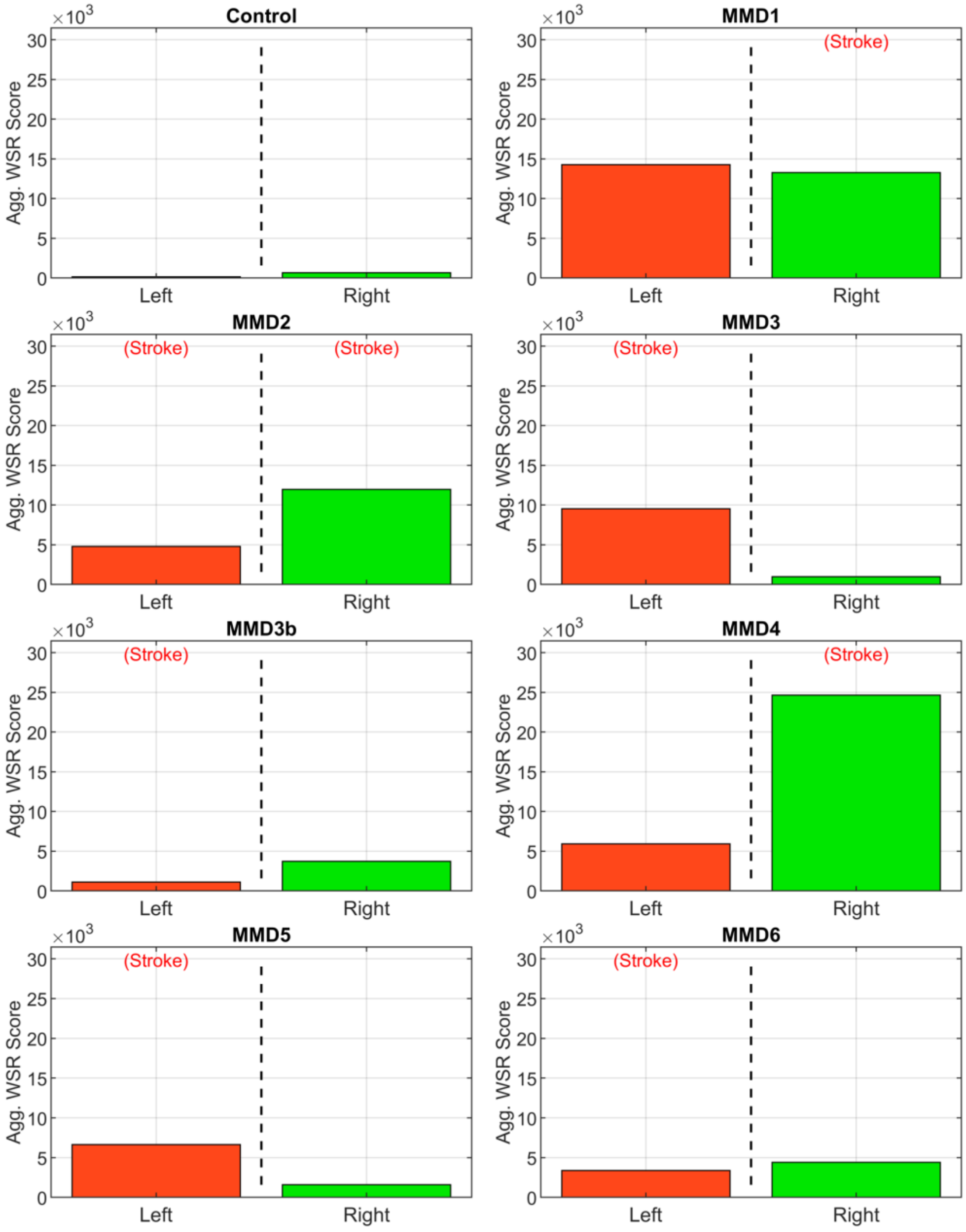


**Supplementary Figure S8.** Aggregate WSR score. Area under the WSR distribution curves from Fig. 4 aggregated by side for the control subject and the 6 patients we studied: MMD1, MMD2, MMD3 and MMD3b, MMD4, MMD5 and MMD6. For each patient, the values shown in Fig. 5 are added. The label (Stroke) indicates the side of primary stroke event.
